# Invasive cervical cancers after an HPV-negative test: insights from screening histories

**DOI:** 10.64898/2026.04.11.26350679

**Authors:** Sadaf Sakina Hassan, Sara Nordqvist Kleppe, Nijin Asinger, Jiangrong Wang, Joakim Dillner, Laila Sara Arroyo Mühr

## Abstract

Human papillomavirus (HPV) testing is the primary method for cervical cancer screening, and a negative HPV test is associated with a very low subsequent risk of invasive cancer. Nevertheless, a small number of cervical cancers are diagnosed following an HPV-negative testing result, posing challenges within HPV-based screening pathways.

Using nationwide Swedish registry data of HPV testing, we identified women diagnosed with invasive cervical cancer between 2019 and 2024 and reconstructed HPV testing histories from the National Cervical Screening Registry (NKCx). The most recent HPV test prior to diagnosis was defined as the index test, and longitudinal HPV testing trajectories were classified among women with an HPV-negative index test.

Of 3,000 women diagnosed with invasive cancer, 243 (8.1%) had an HPV-negative index test. These women were older at diagnosis and more frequently diagnosed at advanced stages compared with women with an HPV-positive index test. Most HPV-negative index tests (66.3%) were performed in the peri-diagnostic period (+/- 30 days). Among women with an HPV-negative index test, 52.7% (128/243) had no prior HPV testing recorded, while the remainder had consistently HPV-negative histories (33.3%, 83/243) or evidence of prior HPV positivity before the index negative test (14%, 32/243). Possible recurrent HPV positivity following an intervening negative test was rare (0.4%, 1/243).

HPV-negative screening results preceding invasive cancer reflect heterogeneous screening histories and cannot be explained solely by test failure. Findings highlighting the importance of reaching women earlier in screening programs and show that fluctuating HPV detectability is rare.

**Novelty and impact:** HPV-negative results preceding cervical cancer are interpreted as test failures, yet underlying screening histories have not been systematically described. Using nationwide registry data, this study reconstructs HPV testing trajectories. HPV-negative cancers occur in older women with limited screening and advanced-stage disease, but also in women with consistently HPV-negative or previously HPV-positive histories. Findings indicate that such cancers cannot be explained solely by test failure and highlight the importance of earlier screening and rare HPV fluctuation.

## INTRODUCTION

Human papillomavirus (HPV) testing has replaced cytology as the primary screening method for cervical cancer in many countries, owing to its high sensitivity for detection of cervical precancer and cancer.^1^ In organized screening programs, a negative HPV test is associated with a very low short- and medium-term risk of invasive cervical cancer, allowing for extended screening intervals and forming a cornerstone of risk-based screening strategies.^2–4^

Despite the high sensitivity of HPV testing, a small number of invasive cervical cancers continue to occur in women with a recent HPV-negative test result.^5,6^ These cancers pose a particular clinical and programmatic challenge, as an HPV-negative result is commonly interpreted as reassurance of low cancer risk and may raise questions regarding test performance and screening pathways. Importantly, an HPV-negative screening test does not imply that the subsequently diagnosed cancer is HPV-negative at the tissue level. Previous studies based on tumor tissue analysis have shown that cervical cancers initially classified as HPV-negative by standard assays may, upon more sensitive or unbiased testing, be reclassified as HPV-positive in a proportion of cases, while tumors remaining HPV-negative represent a distinct subgroup associated with older age at diagnosis, more advanced stage, and poorer prognosis.^7–10^

Several explanations have been proposed for the occurrence of invasive cancer following an HPV-negative test, including sampling or analytical false-negative results, histological subtypes with lower viral detectability and misclassification of tumors of non-cervical origin.^8,11^ Previous studies have quantified the long-term risk of cervical cancer following a negative HPV test;^4,12^ however, little is known about the longitudinal screening histories underlying these cases in real-world screening programs. In particular, it remains unclear whether these HPV-negative results predominantly occur in women with no prior HPV testing, in women with consistently HPV-negative screening histories, or in women with evidence of prior HPV positivity.

Understanding the screening histories that precede invasive cancer diagnoses after an HPV-negative test is essential for interpreting such results within HPV-based screening pathways. Distinguishing between different patterns of HPV test results within the screening history may help contextualize HPV-negative results and inform clinical and programmatic decision-making without inferring biological mechanisms that cannot be assessed using screening data alone. In settings with nationwide cervical screening registries, linkage of longitudinal HPV testing histories with cancer outcomes enables detailed reconstruction of screening trajectories preceding invasive cancer.^13^

In this nationwide registry-based study, we analyzed invasive cervical cancers diagnosed after an HPV-negative test and reconstructed individual HPV testing histories prior to diagnosis. The aim was to characterize longitudinal HPV testing trajectories preceding diagnosis, including the presence or absence of earlier HPV tests, consistency of HPV test results over time, and timing relative to cancer diagnosis, and to determine how HPV-negative results arise within real-world screening contexts.

## MATERIAL AND METHODS

### Study design, data sources, and population

We conducted a nationwide, registry-based observational study to describe human papillomavirus (HPV) testing histories preceding invasive cancer diagnoses. Women diagnosed with an invasive cervical cancer during the period 2019–2024 were identified from the Swedish Gynecological Cancer Registry. Cancer-related variables extracted from the registry included age at diagnosis, FIGO stage, histological subtype, and date of diagnosis. If multiple invasive gynecological cancer diagnoses were recorded for the same woman during 2019–2024, we retained the earliest diagnosis date within the study period as the index cancer.

Cancer registry data were linked to the National Cervical Screening Registry (NKCx; nkcx.se), which contains nationwide records of HPV testing in Sweden, including tests performed within the organized cervical screening program as well as HPV tests performed outside screening, such as opportunistic testing and tests conducted as part of clinical assessment or diagnostic work-up. The registry provides nationwide, virtually complete capture of cervical screening data in Sweden.

For each woman, HPV testing history prior to the cancer diagnosis date was retrieved from NKCx. The index HPV test was defined as the most recent HPV test performed before the cancer diagnosis date or within 30 days of diagnosis. The ±30-day window was used to capture HPV tests performed immediately prior to or during the diagnostic evaluation period, acknowledging that, although the cancer diagnosis date is defined by histopathological confirmation, relevant clinical investigations and HPV testing may occur shortly before this date. When multiple HPV tests were available within this window, the test closest in time to the cancer diagnosis date was selected. HPV tests recorded more than 30 days after the cancer diagnosis were excluded from the analysis.

### Classification of HPV test results and timing

HPV test results were classified according to NKCx coding as HPV positive, HPV negative, invalid (including inadequate or indeterminate samples), or no HPV test recorded prior to diagnosis. Time between the index HPV test and the cancer diagnosis date was calculated and categorized as peri-diagnostic (±30 days) or pre-diagnostic, including intervals of less than one year, one to two years, three to five years, or more than five years prior to diagnosis.

Sensitivity analyses excluded peri-diagnostic HPV tests obtained on or after the diagnosis date, as HPV tests performed in close temporal proximity to diagnosis may be influenced by diagnostic or therapeutic procedures (e.g., biopsy or conization), potentially affecting HPV detection.

### Reconstruction of longitudinal HPV testing histories

For women with an HPV-negative index test, all HPV tests recorded in NKCx prior to the cancer diagnosis were extracted and ordered chronologically. Based on these longitudinal records, individual HPV testing histories were descriptively classified as having no prior HPV testing, consistently HPV-negative results across all recorded tests, or evidence of prior HPV positivity before the HPV-negative index test. The latter category included histories with only HPV-positive results prior to the index negative test as well as histories with mixed HPV-positive and HPV-negative results.

Where available, HPV-positive tests were further classified according to reported genotype information, including HPV16, HPV18, or other high-risk HPV types.

### Statistical analysis

Analyses were descriptive. Characteristics of women and cancers were summarized according to index HPV test status using counts and percentages for categorical variables and medians with interquartile ranges for continuous variables. Group comparisons were performed using chi-square or Fisher’s exact tests for categorical variables and Wilcoxon rank-sum tests for continuous variables, as appropriate. All statistical tests were two-sided, and a p value <0.05 was considered statistically significant. Among women with an HPV-negative index test, the distribution of prior HPV testing histories was summarized. Individual HPV testing trajectories were visualized using a timeline plot in which each woman was represented by a row and each HPV test by a point positioned relative to the cancer diagnosis date. All analyses were conducted using SAS software (SAS Institute Inc., Cary, NC).

### Ethics

The study was conducted in accordance with Swedish ethical and legal requirements for registry-based research. The Regional Ethics Committee in Stockholm, Sweden, granted ethical approval for this study (DNR 2011/1026-31/4, 2023-00289-02, 2025-08540-02) and concluded that informed consent from the study subjects was not required.

## RESULTS

### Study population and index HPV test status

A total of 3,000 women diagnosed with invasive gynecological cancer between 2019 and 2024 were identified from the Swedish Gynecological Cancer Registry and included in the analysis. For 2,405 women (80.2%), at least one HPV test result prior to diagnosis was available in the National Cervical Screening Registry, while 595 women (19.8%) had no recorded HPV test before diagnosis.

Based on the most recent HPV test prior to diagnosis (index HPV test), 2,148 women (71.6%) had an HPV-positive result, 243 women (8.1%) had an HPV-negative result, and 14 women (0.5%) had an invalid or indeterminate result. Women with an HPV-negative index test were significantly older at diagnosis than those with an HPV-positive index test, with a median age of 62 years versus 45 years, respectively (p < 0.001; Table 1).

**Table 1.** Characteristics of invasive gynecological cancers by index HPV test status.

| Characteristic | Total | HPV positive | HPV negative | Other/invalid | No HPV test |
| --- | --- | --- | --- | --- | --- |
| **N** | 3000 | 2148 (71.6%) | 243 (8.1%) | 14 (0.5%) | 595 (19.8%) |
| **Age, median (IQ)** | 50 (28.75) | 45 (23) | 62 (25) | 54.5 (40.25) | 69 (28) |
| **FIGO stage** |  |  |  |  |  |
| **Stage I** | 1722 | 1478 (85.8%) | 86 (5.0%) | 8 (0.5%) | 150 (8.7%) |
| **Stage II** | 977 | 537 (55.0%) | 127 (13.0%) | 4 (0.4%) | 309 (31.6%) |
| **Stage III** | 142 | 70 (49.3%) | 12 (8.5%) | 1 (0.7%) | 59 (41.5%) |
| **Stage IV** | 67 | 20 (29.9%) | 12 (17.9%) | 0 (0.0%) | 35 (52.2%) |
| **Unknown** | 92 | 43 (46.7%) | 6 (6.5%) | 1 (1.1%) | 42 (45.7%) |
| **Histology** |  |  |  |  |  |
| **Squamous** | 2030 | 1446 (71.2%) | 140 (6.9%) | 8 (0.4%) | 436 (21.5%) |
| **Adenocarcinoma** | 862 | 661 (76.7%) | 83 (9.6%) | 3 (0.3%) | 115 (13.3%) |
| **Mixed** | 23 | 20 (87.0%) | 0 (0.0%) | 0 (0.0%) | 3 (13.0%) |
| **Other** | 85 | 21 (24.7%) | 20 (23.5%) | 3 (3.5%) | 41 (48.2%) |
| **Timing of most recent HPV test** |  |  |  |  |  |
| **Peri-dx ($\pm 30$ days)** | 1127 | 956 (84.8%) | 161 (14.3%) | 10 (0.9%) | NA |
| **<1 year** | 886 | 859 (97.0%) | 24 (2.7%) | 3 (0.3%) | NA |
| **1–2 years** | 345 | 308 (89.3%) | 36 (10.4%) | 1 (0.3%) | NA |
| **3–5 years** | 39 | 21 (53.8%) | 18 (46.2%) | 0 (0.0%) | NA |
| **>5 years** | 8 | 4 (50.0%) | 4 (50.0%) | 0 (0.0%) | NA |
| **Women with at least 1 test at and/or prior to diagnosis** | 2405 | 2148 (89.3) | 243 (10.1%) | 14 (0.6%) | NA |

Frequency (percentage) of cervical cancer cases per FIGO stage, histology or timing of most recent HPV-test.
Age is presented as median (interquartile range). Timing of the most recent HPV test is shown among women with at least one recorded HPV test prior to diagnosis. Dx: diagnosis; HPV: human papillomavirus; IQ: interquartile.

### Cancer characteristics by index HPV test result

Cancer stage and histological subtype varied according to index HPV test status (Table 1). Among women with an HPV-negative index test, a significantly larger proportion were diagnosed at stage II or higher compared with those with an HPV-positive index test (p < 0.001). HPV-negative index tests were observed across all FIGO stages but were relatively more frequent among cancers diagnosed at later stages.

With respect to histology, squamous cell carcinoma and adenocarcinoma constituted the majority of cancers in all HPV test groups. Among women with an HPV-negative index test, the relative proportion of adenocarcinoma was significantly higher than among those with an HPV-positive index test (p < 0.001). In addition, a significantly higher proportion of non-squamous, non-adenocarcinoma histologies was observed in this group (p < 0.001).

### Timing of the index HPV test relative to cancer diagnosis

The timing of the index HPV test relative to cancer diagnosis was different between HPV-positive and HPV-negative index tests (Table 1). Among women with an HPV-negative index test, 161 (66.3%) had their test performed in the peri-diagnostic period (±30 days), whereas 82 women (33.7%) had an HPV-negative test recorded more than 30 days prior to diagnosis. Of these, 24 tests (29.3%) were performed within one year before diagnosis, 36 tests (43.9%) one to two years before diagnosis, 18 tests (7.4%) three to five years before diagnosis, and four tests (1.6%) more than five years prior to diagnosis. The proportion of cancers diagnosed after an HPV-negative index test was lowest within one year of diagnosis (2.7%) and increased in longer time intervals. However, estimates at longer intervals were based on small numbers of cases and should be interpreted with caution.

### Longitudinal HPV testing histories among women with an HPV-negative index test

Among the 243 women with an HPV-negative index test, 128 women (52.7%) had no prior HPV test recorded in NKCx before the index test. The remaining 115 women (47.3%) had at least one HPV test recorded before the HPV-negative index test. Individual HPV testing trajectories for women with an HPV-negative index test are shown in Figure 1.

**Figure 1.**
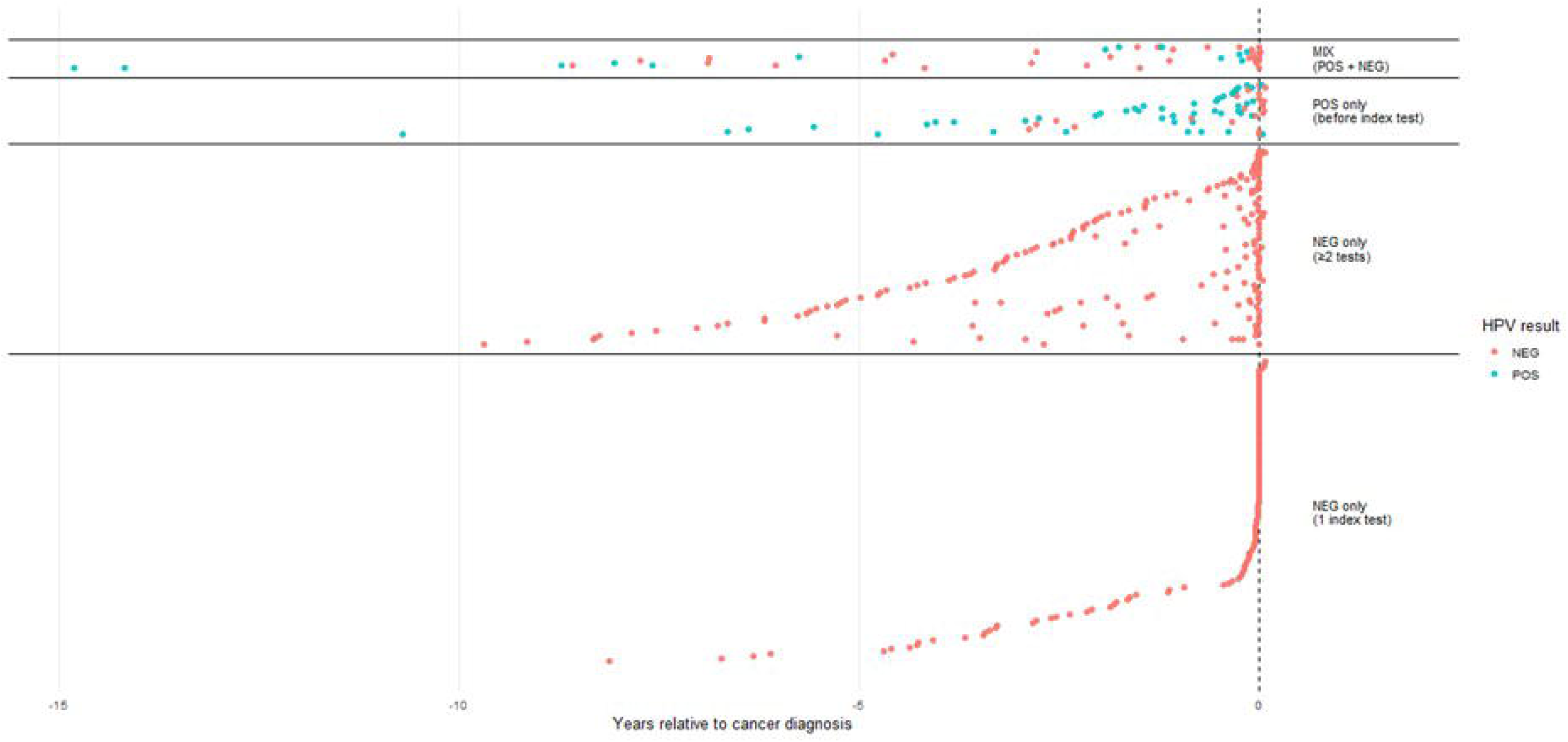
Individual HPV testing trajectories for women with an HPV-negative index test before cancer diagnosis. Each row represents one woman, and each point represents an HPV test result plotted relative to the date of cancer diagnosis (year 0, dashed vertical line). Red points indicate HPV-negative results and blue points indicate HPV-positive results. Women are grouped according to their longitudinal HPV testing history prior to the HPV-negative index test: no prior HPV testing (NEG only (1 index test)), consistently HPV-negative testing histories (NEG only (>= 2 tests), HPV-positive tests only before the index negative test (POS only (before the index test), and mixed HPV-positive and HPV-negative testing histories (MIX (POS+NEG)).

Of the 115 women with prior HPV testing, 83 women (34.2% of total women with an HPV-negative index test) had consistently HPV-negative results across all recorded tests prior to diagnosis. These women had a median of two HPV tests before diagnosis (range, 2–6). In contrast, 32 women (13.2% of total women with an HPV-negative index test) had evidence of HPV positivity before the HPV-negative index test and had a median of three HPV tests before diagnosis (range, 2–7). Among these, 22 women had only HPV-positive test results prior to the index negative test, while 10 women (4.1% of total women with an HPV-negative index test) had a mixture of HPV-positive and HPV-negative results prior to the index test. Detailed review of individual testing histories showed that among women with mixed histories, nearly all women (9/10) either transitioned from HPV-positive to persistently HPV-negative results or had isolated HPV-positive tests that did not recur. Only one woman showed a pattern suggestive of recurrent HPV positivity, with a high-risk HPV (non-16/18) positive result, followed by a negative test, a subsequent positive result, and then two consecutive negative tests (Figure 2).

**Figure 2.**
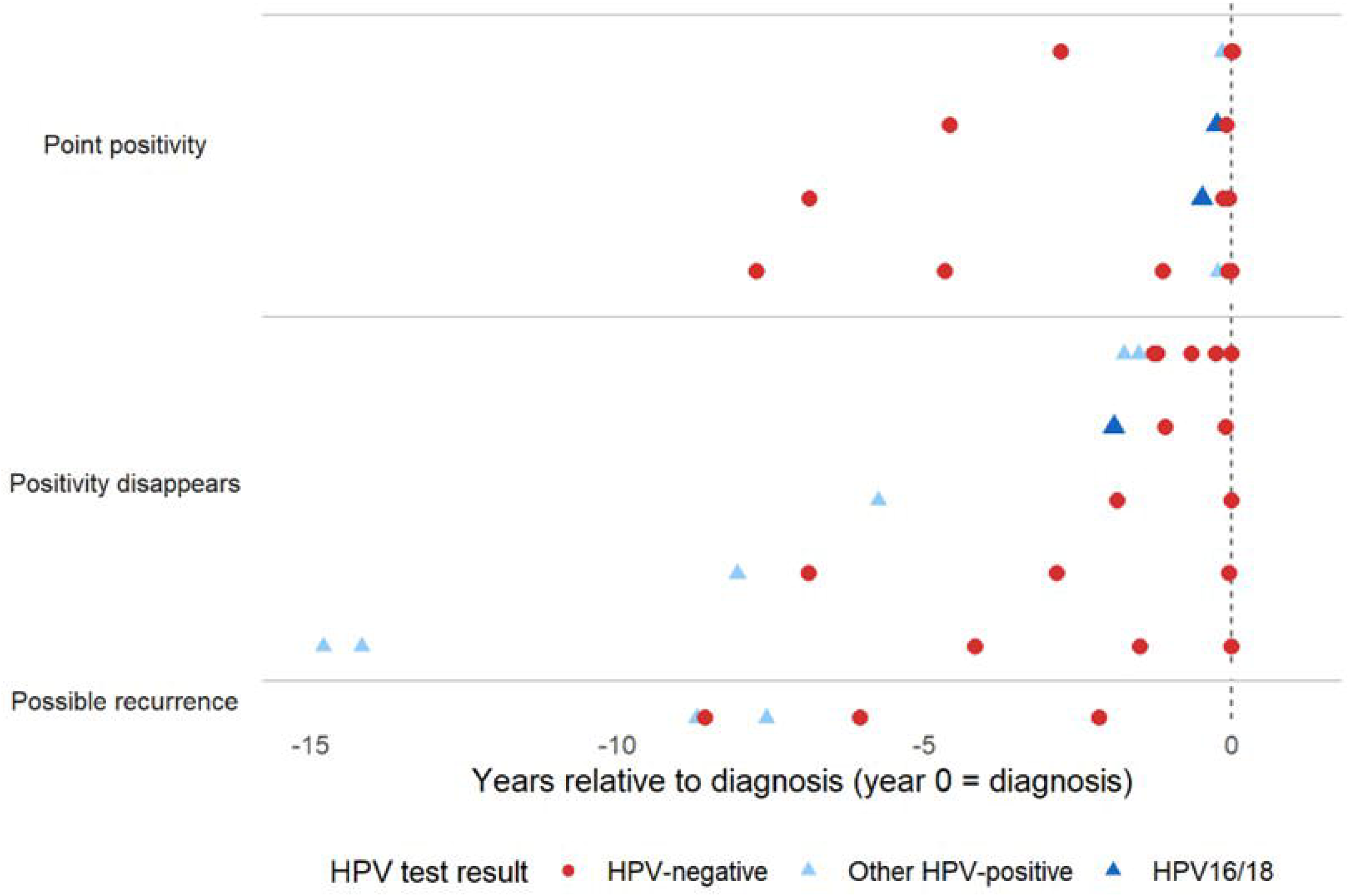
Longitudinal HPV test results among women with mixed HPV histories prior to the index test The figure includes only women with a mixed HPV testing history defined as having both HPV-positive and HPV-negative results prior to the index test (n=10). Each row represents an individual woman. Colored symbols indicate HPV test results at the corresponding time relative to cervical cancer diagnosis (year 0). Red symbols denote HPV-negative results, light blue symbols denote positivity for other high-risk HPV types, and dark blue symbols denote HPV16/18 positivity. Women are grouped according to their longitudinal HPV testing patterns. Time is shown as years relative to diagnosis.

In sensitivity analyses excluding peri-diagnostic HPV tests obtained on or after the diagnosis date, a total of 181 women had an HPV-negative index test prior to diagnosis. Among these, 88 (48.61%) had no prior HPV testing recorded, 66 (36.46%) had consistently HPV-negative testing histories (median 2 tests per woman, maximum 6), 18 (9.94%) had only HPV-positive results prior to the index negative test, and 9 (4.97%) had mixed HPV-positive and HPV-negative results. Excluding peri-diagnostic tests did not materially alter the observed history patterns.

## DISCUSSION

In this nationwide registry-based study, we identified and characterized invasive cervical cancers diagnosed over a six-year period according to preceding HPV testing and examined the longitudinal HPV testing histories of women with an HPV-negative index test.

Approximately 8% of cancers were preceded by an HPV-negative index test. These women were older at diagnosis and more frequently diagnosed at advanced stages compared with women with an HPV-positive index test, highlighting distinct clinical and epidemiological features associated with HPV-negative screening results. These findings are in line with previous studies based on tumor HPV status, which have shown that HPV-negative cervical cancers are more often diagnosed at older ages, at more advanced stages, and are associated with poorer prognosis.^9^

The predominance of peri-diagnostic HPV-negative tests, together with the older age and more advanced stage at diagnosis, is consistent with many HPV-negative index tests being performed at the time of cancer diagnosis, potentially as part of clinical assessment rather than routine screening. This distinction is relevant for screening program evaluation and quality assurance, as peri-diagnostic testing may not reflect true screening performance.

In addition to differences in age, timing, and stage, histological patterns also varied by index HPV test status. Women with an HPV-negative index test had a relatively higher proportion of adenocarcinoma and non-squamous, non-adenocarcinoma histologies compared with women with an HPV-positive index test. These findings are consistent with previous reports indicating lower HPV detectability in certain histological subtypes and underscore the importance of considering tumor characteristics when interpreting HPV-negative screening results.^9,14^

Longitudinal reconstruction of HPV testing histories demonstrated that HPV-negative index tests reflected heterogeneous prior screening trajectories rather than a single underlying pattern. More than half of women with an HPV-negative index test had no prior HPV testing recorded, indicating that a substantial proportion of HPV-negative results occurred in women without an established HPV testing history. HPV tests obtained in close temporal proximity to diagnosis may be influenced by diagnostic or therapeutic procedures, potentially affecting HPV detection. However, exclusion of peri-diagnostic tests performed on the date and/or after the diagnosis did not materially alter the observed distribution of longitudinal HPV testing histories.

Among women with repeated HPV testing, most had consistently HPV-negative results over time, while a smaller subset had evidence of prior HPV positivity before the index negative test. The predominance of consistently HPV-negative testing histories suggests that repeated testing over short time intervals is unlikely to substantially alter HPV detection patterns in this subgroup.

Possible recurrent HPV positivity following an intervening negative test was rare (1/3000), indicating that fluctuating detectability of HPV infection is uncommon when high-quality testing is applied in a real-world screening program. This finding supports the robustness and reliability of HPV-based screening.

Although this observation may reflect multiple factors, including variability in viral detectability, sampling, or lesion evolution, these mechanisms cannot be resolved using screening registry data alone. Therefore, complementary measures to ensure high-quality HPV testing remain essential, including regular laboratory audits, systematic review of cervical cancer cases, and re-testing of HPV-negative high-grade lesions.^11,15,16^

These findings help contextualize the longstanding discussion on HPV latency and recurrent detectability. While intermittent HPV positivity after a negative test has been described, such patterns were exceedingly rare in our material. This is consistent with large-scale screening studies showing that a single negative HPV test confers long-term protection against cervical cancer.^3,4,17^ If clinically meaningful latent or frequently recurring HPV infections after a negative test were common, the well-established long-term reassurance associated with a negative HPV result would not be observed. Thus, although latency cannot be excluded at the individual level, our findings suggest that it has limited relevance for population-based screening performance.

The subgroup of women with consistently HPV-negative testing histories is also informative for understanding the performance limits of HPV-based screening. In this study, cancers occurring in women with only HPV-negative tests prior to diagnosis represented approximately 3% of all cases (83/3000), suggesting that the theoretical maximum sensitivity of HPV testing in routine practice may be close to 97%. Importantly, these cases are unlikely to be prevented by shortening screening intervals, as repeated testing does not appear to increase detection in this subgroup. Instead, they likely reflect the upper boundary of test performance in real-world conditions, influenced by factors such as sampling limitations, tumor characteristics, or cancers with low or absent HPV detectability. These findings suggest that, in many cases, cancers following an HPV-negative test reflect delayed entry into effective screening rather than failure of repeated surveillance. The combination of limited prior screening, older age, and more advanced stage at diagnosis indicates that many of these cancers arise in women who were not reached early enough by screening programs. In this respect, these cases share important characteristics with cancers occurring in non-attenders, where the primary limitation is not screening frequency but failure to detect disease at an earlier, more treatable stage.

Strengths of this study include its nationwide coverage, use of high-quality population-based registries with virtually complete capture of HPV testing and cancer diagnoses, and the ability to link individual-level screening histories to cancer outcomes. These features enabled both characterization of cancer features by HPV test status and detailed reconstruction of prior testing trajectories in a real-world screening setting.

Several limitations should be acknowledged. HPV negativity in this study refers to the screening test result rather than the biological HPV status of the cancer. In addition, the number of women with mixed HPV-positive and HPV-negative testing histories was small, limiting more detailed subgroup analyses. Furthermore, a proportion of women had no recorded HPV test prior to diagnosis. The proportion of women without prior HPV testing (595/3000, 19.8%) decreased over calendar time, reflecting the gradual implementation and increasing coverage of HPV-based screening in Sweden. Given the nationwide and virtually complete capture of cervical screening data in the registry, the absence of prior HPV testing reflects true lack of screening rather than incomplete data capture.

In conclusion, cancers diagnosed after an HPV-negative screening test represent a distinct subgroup characterized by older age, more advanced stage, and heterogeneous prior HPV testing histories. HPV-negative index tests preceding invasive cancer arise primarily in four screening-related patterns: absence of prior HPV testing, consistently HPV-negative testing histories, prior HPV positivity followed by repeated HPV-negative test results before diagnosis, and, very rarely, mixed HPV-positive and HPV-negative testing histories. These findings underscore the importance of interpreting HPV-negative screening results in the context of longitudinal testing histories and tumor characteristics when evaluating cancers occurring after a negative HPV screen.

## AUTHOR CONTRIBUTIONS

Conceptualization and project administration: LSAM. Resources: LSAM, JD. Methodology, investigation, and validation: LSAM, SSH, NA, JW, SNK. Formal analysis and data curation: LSAM, SSH, NA. Writing – original draft: LSAM. Writing – review and editing: SSH, NA, JW, SNK, JD. All authors critically reviewed the manuscript, approved the final version, and take responsibility for the integrity of the work and the decision to submit the manuscript for publication.

## Abbreviations

CIN3+: cervical intraepithelial neoplasia grade 3 or worse
Dx: diagnosis
HPV: human papillomavirus
NKCx: National Cervical Screening Registry
FIGO: International Federation of Gynecology and Obstetrics
IQR: interquartile range

## Data Availability

The data used in this study were obtained from national Swedish health registries. Access to these data is subject to approval by the relevant registry holders and ethical review authorities, in accordance with Swedish law and data protection regulations.
Researchers interested in accessing data from the National Cervical Screening Registry (NKCx; nkcx.se) and the Swedish Gynecological Cancer Registry may apply directly to the respective registry custodians after obtaining appropriate ethical approval and completing the required data access agreements.
Further information about the data access process is available from the corresponding author upon reasonable request.

## REFERENCES

1. Ronco G, Dillner J, Elfström KM, et al. Efficacy of HPV-based screening for prevention of invasive cervical cancer: follow-up of four European randomised controlled trials. The Lancet. 2014;383(9916):524–532. doi:10.1016/S0140-6736(13)62218-7

2. Dillner J, Gray P. [HPV screening coverage and cancer]. Lakartidningen. 2025;122:25007.

3. Elfstrom KM, Smelov V, Johansson ALV, et al. Long term duration of protective effect for HPV negative women: follow-up of primary HPV screening randomised controlled trial. BMJ. 2014;348(jan16 1):g130–g130. doi:10.1136/bmj.g130

4. Wang J, Elfström KM, Dillner J. Human papillomavirus-based cervical screening and long-term cervical cancer risk: a randomised health-care policy trial in Sweden. Lancet Public Health. 2024;9(11):e886–e895. doi:10.1016/S2468-2667(24)00218-4

5. Lagheden C, Eklund C, Lamin H, et al. Nationwide comprehensive human papillomavirus (HPV) genotyping of invasive cervical cancer. Br J Cancer. 2018;118(10):1377–1381. doi:10.1038/s41416-018-0053-6

6. Tjalma W. HPV negative cervical cancers and primary HPV screening. Facts Views Vis ObGyn. 2018;10(2):107–113.

7. Arbyn M, Verdoodt F, Snijders PJF, et al. Accuracy of human papillomavirus testing on self-collected versus clinician-collected samples: a meta-analysis. Lancet Oncol. 2014;15(2):172–183. doi:10.1016/S1470-2045(13)70570-9

8. Arroyo Mühr LS, Lagheden C, Lei J, et al. Deep sequencing detects human papillomavirus (HPV) in cervical cancers negative for HPV by PCR. Br J Cancer. 2020;123(12):1790–1795. doi:10.1038/s41416-020-01111-0

9. Lei J, Arroyo-Mühr LS, Lagheden C, et al. Human Papillomavirus Infection Determines Prognosis in Cervical Cancer. J Clin Oncol. 2022;40(14):1522–1528. doi:10.1200/JCO.21.01930

10. Lei J, Ploner A, Lagheden C, et al. High-risk human papillomavirus status and prognosis in invasive cervical cancer: A nationwide cohort study. Zheng W, ed. PLOS Med. 2018;15(10):e1002666. doi:10.1371/journal.pmed.1002666

11. Prétet JL, Arroyo Mühr LS, Cuschieri K, et al. Human papillomavirus negative high grade cervical lesions and cancers: Suggested guidance for HPV testing quality assurance. J Clin Virol. 2024;171:105657. doi:10.1016/j.jcv.2024.105657

12. Gilham C, Sargent A, Crosbie EJ, Peto J. Long-term risks of invasive cervical cancer following HPV infection: follow-up of two screening cohorts in Manchester. Br J Cancer. 2023;128(10):1933–1940. doi:10.1038/s41416-023-02227-9

13. Arroyo Mühr LS, Wang J, Hassan SS, Yilmaz E, Elfström MK, Dillner J. Nationwide registry-based trial of risk-stratified cervical screening. Int J Cancer. 2025;156(2):379–388. doi:10.1002/ijc.35142

14. Giannella L, Di Giuseppe J, Delli Carpini G, et al. HPV-Negative Adenocarcinomas of the Uterine Cervix: From Molecular Characterization to Clinical Implications. Int J Mol Sci. 2022;23(23):15022. doi:10.3390/ijms232315022

15. Hortlund M, Mühr LSA, Lagheden C, Hjerpe A, Dillner J. Audit of laboratory sensitivity of human papillomavirus and cytology testing in a cervical screening program. Int J Cancer. 2021;149(12):2083–2090. doi:10.1002/ijc.33769

16. Wang J, Elfström KM, Andrae B, et al. Cervical cancer case–control audit: Results from routine evaluation of a nationwide cervical screening program. Int J Cancer. 2020;146(5):1230–1240. doi:10.1002/ijc.32416

17. Katki HA, Schiffman M, Castle PE, et al. Five-Year Risks of CIN 3+ and Cervical Cancer Among Women With HPV-Positive and HPV-Negative High-Grade Pap Results. J Low Genit Tract Dis. 2013;17(Supplement 1):S50–S55. doi:10.1097/LGT.0b013e3182854282

